# Comparative efficacy of tocilizumab and baricitinib in COVID-19 treatment: a retrospective cohort study

**DOI:** 10.1101/2021.12.13.21267717

**Authors:** Yuichi Kojima, Sho Nakakubo, Keisuke Kamada, Yu Yamashita, Nozomu Takei, Junichi Nakamura, Munehiro Matsumoto, Hiroshi Horii, Kazuki Sato, Hideki Shima, Masaru Suzuki, Satoshi Konno

## Abstract

**Background:** Although biological agents, tocilizumab and baricitinib, have been shown to improve the outcomes of patients with COVID-19, a comparative evaluation has not been performed.

**Methods:** A retrospective, single-center study was conducted using the data of patients with COVID-19 admitted to the Hokkaido University hospital between April 2020 and September 2021, who were treated with tocilizumab or baricitinib. The clinical characteristics of patients who received each drug were compared. Univariate and multivariate logistic regression models were performed against the outcomes of all-cause mortality and the improvement in respiratory status. The development of secondary infection events was analyzed using the Kaplan–Meier analysis and the log-rank test.

**Results:** The use of tocilizumab or baricitinib was not associated with all-cause mortality and the improvement in respiratory status within 28 days of drug administration. Age, chronic renal disease, and comorbid respiratory disease were independent prognostic factors for all-cause mortality, while anti-viral drug use and severity of COVID-19 at baseline were associated with the improvement in respiratory status. There was no significant difference in the infection-free survival between patients treated with tocilizumab and those with baricitinib.

**Conclusion:** There were no differences in efficacy and safety between tocilizumab and baricitinib for the treatment of COVID-19.

## Introduction

Coronavirus disease 2019 (COVID-19), caused by severe acute respiratory syndrome coronavirus 2 (SARS-CoV-2), continues to spread worldwide. As various treatment methods have been established and vaccination has progressed, the situation surrounding COVID-19 has entered a new phase. However, the optimal treatment for severe COVID-19 continues to be explored.

Steroids were first identified to be effective in the treatment of severe COVID-19 and are now considered as the standard treatment (1, 2). Following this, several studies were conducted to determine whether addition of biological agents to the standard treatment could improve prognosis. Tocilizumab (TCZ) is a monoclonal antibody against interleukin-6 receptor-alpha (3). Both REMAP-CAP and RECOVERY trials evaluated the add-on effect of TCZ to the standard of care in hospitalized patients with severe-to-critical COVID-19 and showed that TCZ reduces mortality or prolongs organ support-free days (4, 5). Baricitinib (BRT) is a Janus kinase (JAK) inhibitor with high selectivity for JAK1 and JAK2 molecules of the JAK family (6). It has been observed initially in the ACTT-2 trial that BRT shortens the time to recovery, when used in combination with remdesivir in the treatment of severe COVID-19 (7). Further, in the COV-BARRIER trial, treatment with baricitinib, in addition to standard care, was associated with reduced mortality in adults hospitalized with COVID-19 (8). A common finding among these trials was that both TCZ and BRT are particularly effective in reducing mortality in patients with a high demand for oxygen.

Based on the evidence, the current guidelines recommend that both TCZ and BRT be administered in combination with steroids to patients with severe COVID-19 requiring high-flow oxygen and non-invasive mechanical ventilation and those with rapidly increasing oxygen needs and systemic inflammation (9, 10). However, to our knowledge, no comparative study exists to verify the superiority of the efficacy of TCZ versus BRT against COVID-19; therefore, the priority among the two is not clearly stated in the current international recommendation (9).

Herein, we retrospectively analyzed the medical information of patients with COVID-19 admitted to the Hokkaido University Hospital to compare whether the use of TCZ or BRT was associated with mortality, clinical improvement, and incidence of secondary infection.

## Patients and Methods

### Patients

This single-center, retrospective cohort study was approved by Hokkaido University Hospital Division of Clinical Research Administration(Research No. 020-0107). The requirement for obtaining informed consent was waived by the relevant ethics committee due to the retrospective nature of the study. This study included patients with COVID-19 who were admitted to the Hokkaido University Hospital between April 2020 and September 2021. All patients were confirmed to be positive for SARS-CoV-2 by polymerase chain reaction (PCR). Among the patients, those treated with biological agents (TCZ or BRT) for COVID-19 were selected for the present analysis. Cases in which both drugs were administered during the course of treatment were excluded.

### Data collection

The clinical data (age, sex, body mass index, history of smoking, history of vaccination, comorbidities, respiratory status and severity, days from the onset of COVID-19, treatment protocol laboratory data, and clinical outcome) were collected from medical records. We defined the severity of COVID-19 as follows: severity level 1- hospitalized but not requiring supplemental oxygen; level 2- hospitalized and requiring supplemental oxygen ≤ 4 L per minute (L/min); severity level 3- hospitalized and requiring oxygen therapy ≥ 5 L/min, including receiving nasal high-flow oxygen therapy, non-rebreather, or noninvasive mechanical ventilation; level 4- receiving invasive mechanical ventilation at administration. The reason for setting this severity classification was that in our hospital, the criteria for administering biological agents was when the oxygen administration rate deteriorated to 5 L/min or higher.

We set the following clinical endpoints: all-cause mortality, improvement in respiratory status, and development of secondary infection events within 28 days after administration of TCZ or BRT. Improvement in respiratory status was defined as a recovery in severity to level 1 or 2 after the initiation of TCZ or BRT. Secondary infection events included pneumonia, bacteremia, urinary tract infection, and fungal infection requiring antibiotic treatment,

### Statistical analysis

Continuous data are expressed as median and interquartile range (IQR). Categorical data are expressed as absolute number and percentages. Wilcoxon rank sum test or Kruskal–Wallis test was used to compare differences in continuous variables, and chi-square test or Fisher’s exact test was used to evaluate differences between categorical variables. Clinical outcomes including all-cause mortality, improvement in respiratory status, and development of secondary infection events, were analyzed using univariable and multivariate logistic regression models, with odds ratio (OR) and 95 % confidence intervals (CI). Variables with P < 0.1 in the univariate analysis were entered into the multivariate models. Infection-free survival was evaluated by the Kaplan–Meier method with a log-rank test. All P-values were two-tailed, with statistical significance set at P < 0.05. JMP (SAS Institute Inc., Cary, NC, USA) were used for all statistical processing.

## Results

### Study population

A total of 459 patients were admitted to the Hokkaido University Hospital with the diagnosis of COVID-19 during the study period. Of these, 100 patients received biological agents for treating the symptoms of COVID-19. Sixty-four patients were treated with TCZ (TCZ group) and 34 with BRT (BRT group). Two patients were initially treated with BRT but switched to TCZ, who were excluded from the study (Figure 1).

**Figure 1.**
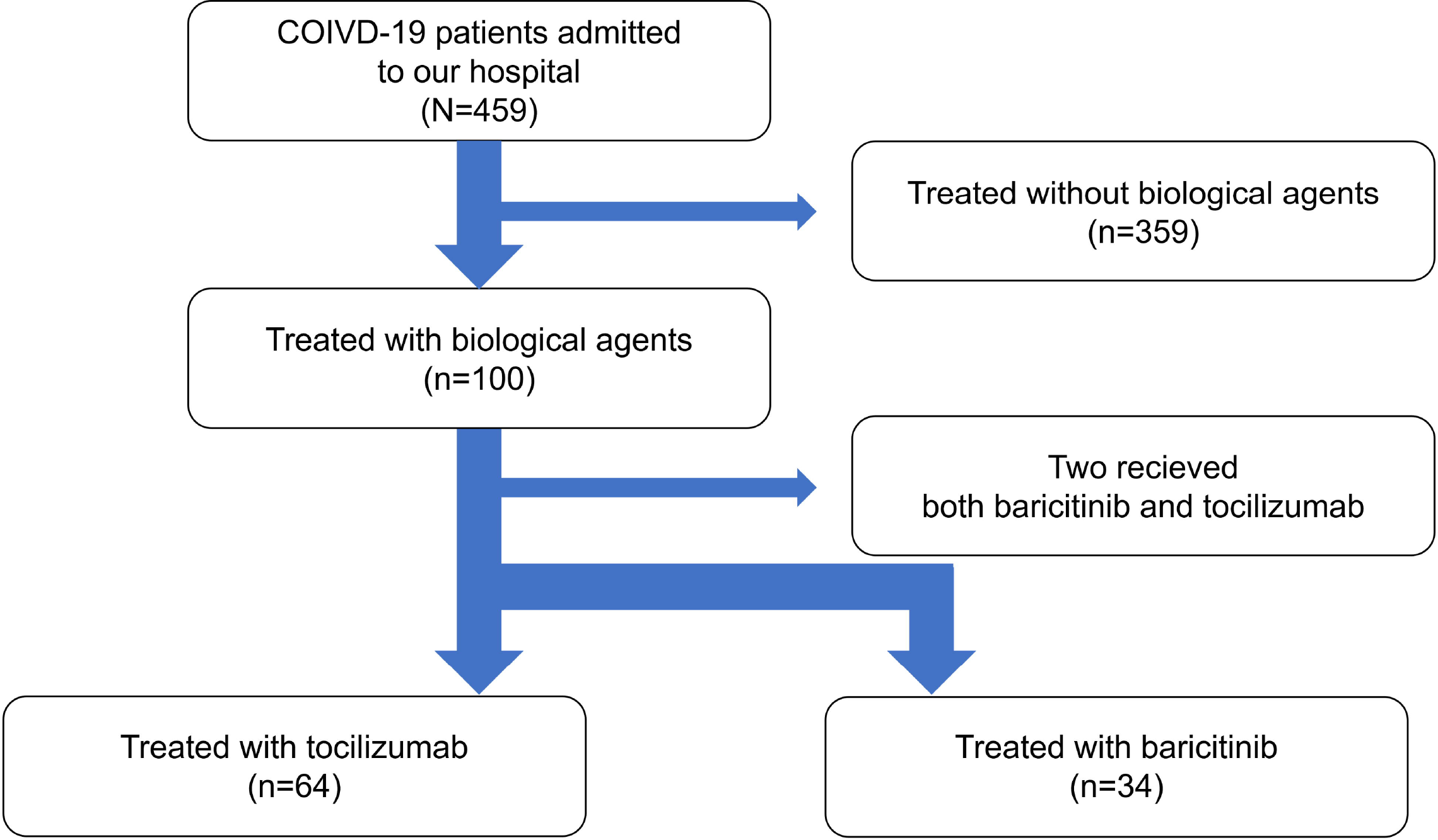
Flow chart of patients with COVID-19 with the tocilizumab and baricitinib groups.

### Baseline characteristics

The median age of total patients (N=98) was 60.5 years, and 74.5 % were males (Table 1). Compared with the TCZ group (n=64), BRT group (n=34) had lower age (58.5 vs. 65.5 years, P=0.03) and lower prevalence of chronic heart disease (5.9 % vs. 23.4 %, P=0.03). There were no significant differences in sex, smoking history, immunosuppressive drug use, obesity, chronic kidney disease, diabetes mellitus, collagen disease, hypertension, and comorbid respiratory disease. Only one patient in BRT group was fully vaccinated with two doses of the vaccine against SARS-CoV-2. Days from the onset of illness to the administration of biological agents were not significantly different between the two groups (10 vs. 9 days, P=0.50). Analysis of blood samples revealed that, compared to the BRT group, the TCZ group had a significantly lower eosinophil count and hemoglobin (0 vs. 0, P=0.047, 13.8 vs. 14.5, P=0.04, respectively), higher levels of lactate dehydrogenase (LDH), krebs von den Lungen-6 (KL-6), and D-dimer (540 vs. 470, P=0.02, 444 vs. 319, P= 0.03, 1.5 vs. 1.0, P<0.01, respectively).

**Table 1.**
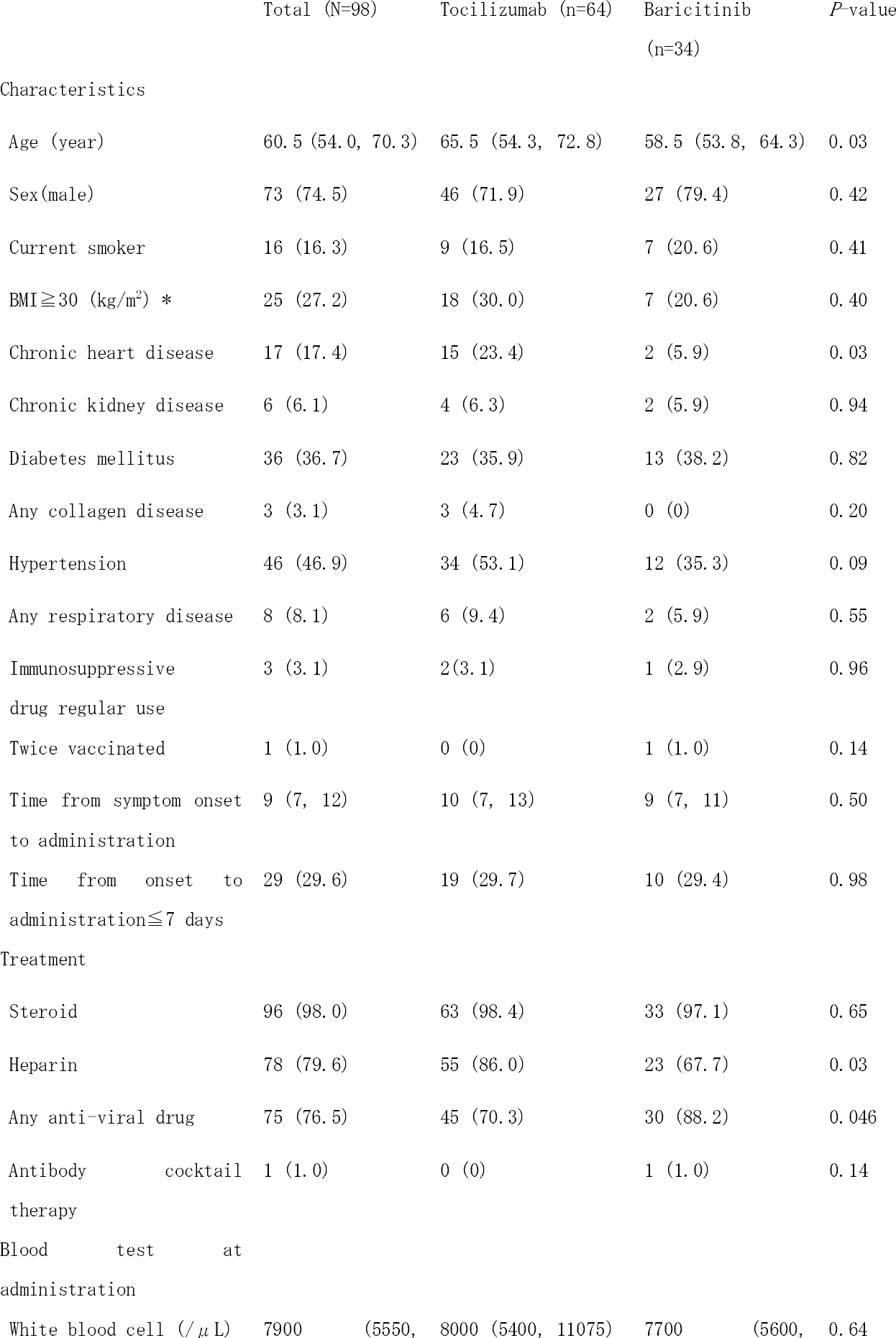
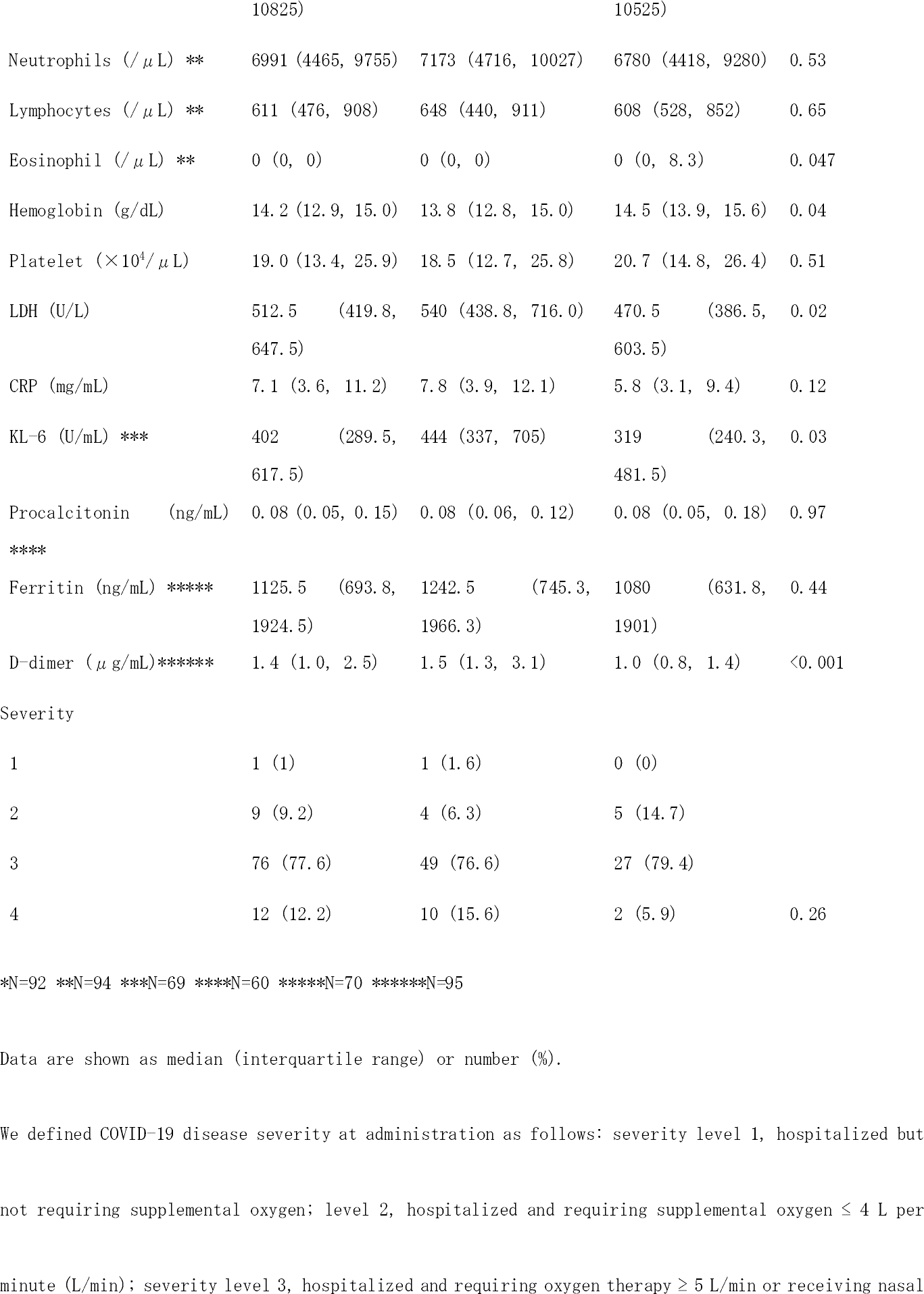
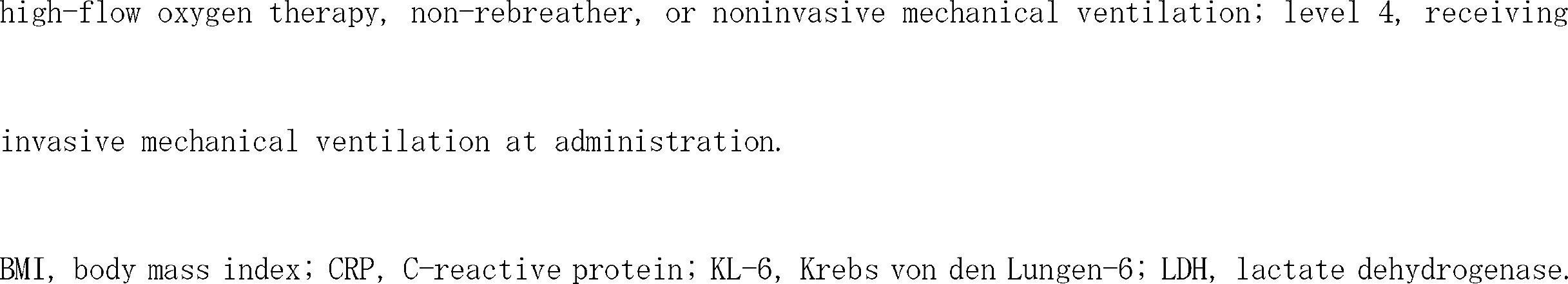
Comparison of baseline characteristics between tocilizumab and baricitinib groups.

Most of the patients in both the groups received steroid treatment for COVID-19 (98.4 % in TCZ and 97.1 % in BRT group). The TCZ group were administered heparin more frequently and antivirals less frequently than the BRT group (86.0 % vs. 67.7 %, P=0.03, 70.3 % vs. 88.2 %, P=0.046, respectively). Only one patient was treated with a combination of the monoclonal antibodies (casirivimab and imdevimab) in the BRT group. The severity of COVID-19 was similar in both groups at the time of initiating treatment with the biological agents, with severity level 3 or higher in 93.2 % of the patients in TCZ group and 85.3 % in BRT group.

### Risk factors for death within 28 days after initiating treatment with biological agents

Among the group of patients administered biological agents (N=98), univariate analysis showed that the use of TCZ was significantly associated with increased all-cause mortality. Additionally, increased age, presence of chronic kidney disease, administration of biological agents at less than seven days from onset, and no antiviral drug use were significantly associated with all-cause mortality (Table 2). In multivariate analysis, older age (OR =1.10, 95 % confidence interval (CI) 1.00 –1.21, P=0.02), presence of chronic kidney disease (OR = 43.10, 95 %CI 2.71 – 686.04, P=0.008), and early administration of biological agents from onset (OR = 18.09, 95 %CI 1.70 –192.47, P=0.02) were shown to be independent risk factors for all-cause mortality within 28 days. In contrast, the use of TCZ was not an independent prognostic factor for death (OR = 13.28, 95 %CI 0.45 – 392.92, P=0.13) (Table 2).

**Table 2.**
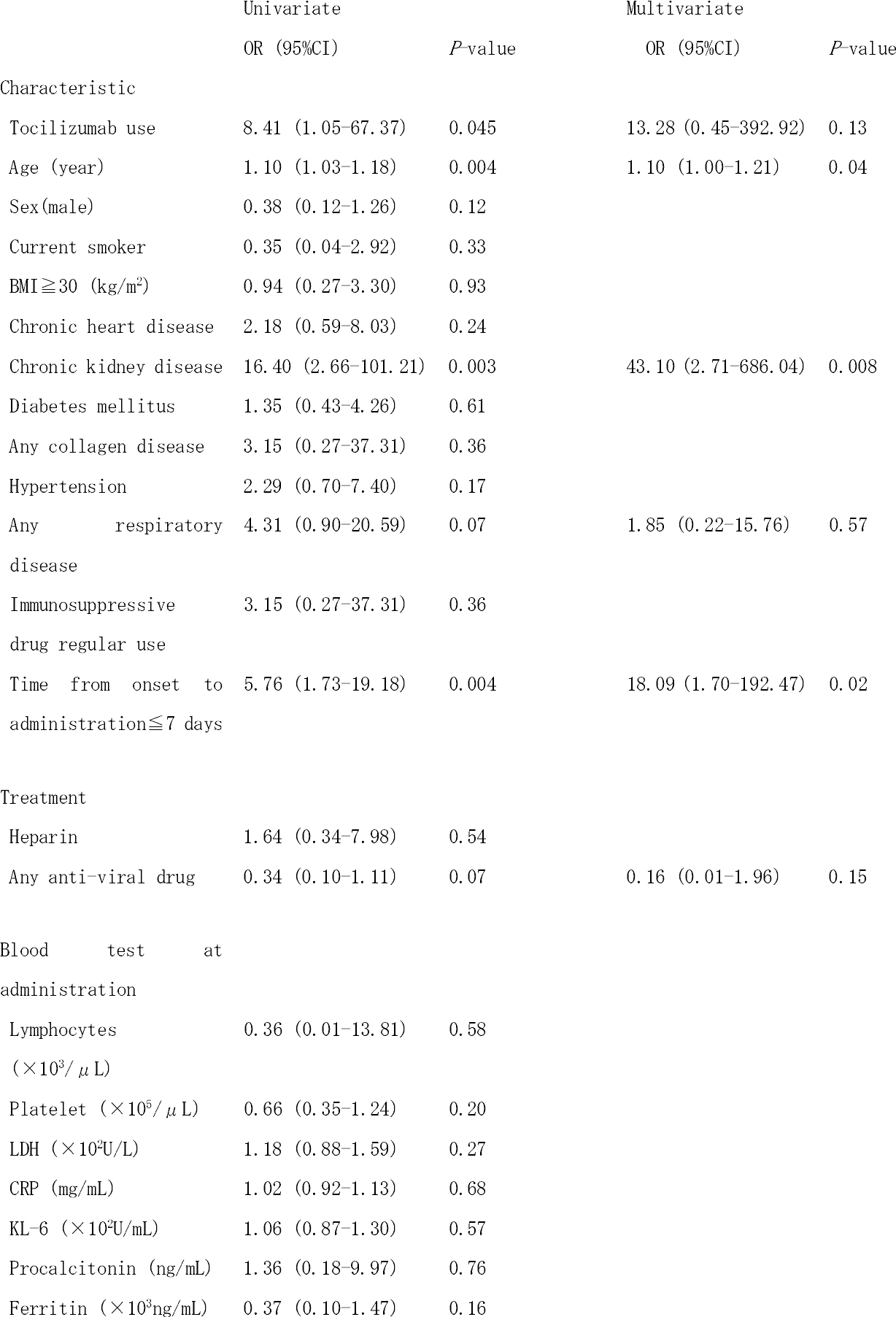
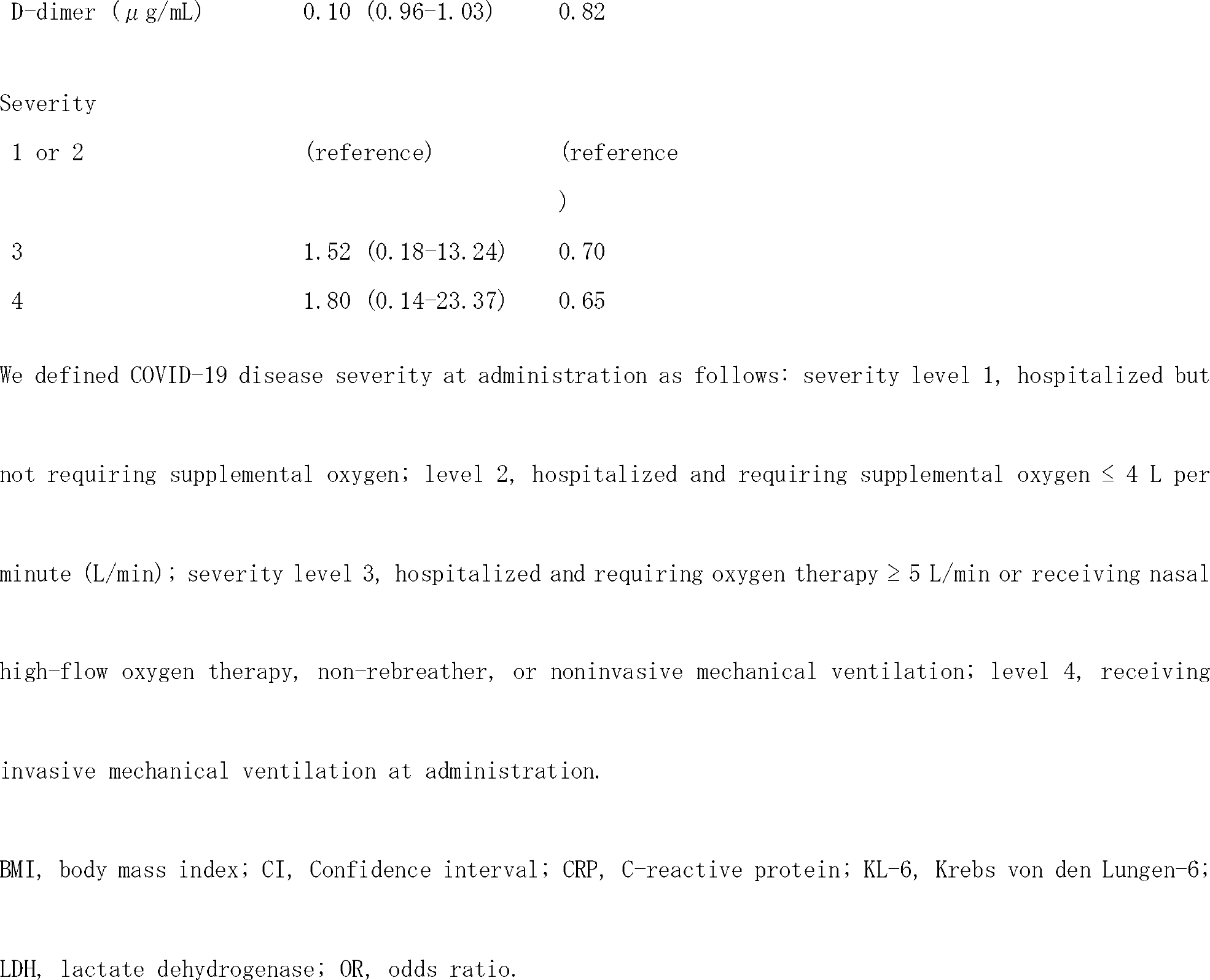
Predicting factors for death within 28 days of administration in patients treated with tocilizumab or baricitinib assessed using logistic regression analysis.

| Characteristic | Univariate |  | Multivariate |  |
| --- | --- | --- | --- | --- |
|  | OR (95%CI) | *P*-value | OR (95%CI) | *P*-value |
| Tocilizumab use | 8.41 (1.05-67.37) | 0.045 | 13.28 (0.45-392.92) | 0.13 |
| Age (year) | 1.10 (1.03-1.18) | 0.004 | 1.10 (1.00-1.21) | 0.04 |
| Sex(male) | 0.38 (0.12-1.26) | 0.12 |  |  |
| Current smoker | 0.35 (0.04-2.92) | 0.33 |  |  |
| BMI≥30 (kg/m<sup>2</sup>) | 0.94 (0.27-3.30) | 0.93 | | |
| Chronic heart disease | 2.18 (0.59-8.03) | 0.24 |  |  |
| Chronic kidney disease | 16.40 (2.66-101.21) | 0.003 | 43.10 (2.71-686.04) | 0.008 |
| Diabetes mellitus | 1.35 (0.43-4.26) | 0.61 |  |  |
| Any collagen disease | 3.15 (0.27-37.31) | 0.36 |  |  |
| Hypertension | 2.29 (0.70-7.40) | 0.17 |  |  |
| Any respiratory disease | 4.31 (0.90-20.59) | 0.07 | 1.85 (0.22-15.76) | 0.57 |
| Immunosuppressive drug regular use | 3.15 (0.27-37.31) | 0.36 |  |  |
| Time from onset to administration≤7 days | 5.76 (1.73-19.18) | 0.004 | 18.09 (1.70-192.47) | 0.02 |
| Treatment |  |  |  |  |
| Heparin | 1.64 (0.34-7.98) | 0.54 |  |  |
| Any anti-viral drug | 0.34 (0.10-1.11) | 0.07 | 0.16 (0.01-1.96) | 0.15 |
| Blood test at administration |  |  |  |  |
| Lymphocytes ($\times 10^3/\mu$ L) | 0.36 (0.01-13.81) | 0.58 | | |
| Platelet ($\times 10^5/\mu$ L) | 0.66 (0.35-1.24) | 0.20 | | |
| LDH ($\times 10^2$U/L) | 1.18 (0.88-1.59) | 0.27 | | |
| CRP (mg/mL) | 1.02 (0.92-1.13) | 0.68 |  |  |
| KL-6 ($\times 10^2$U/mL) | 1.06 (0.87-1.30) | 0.57 | | |
| Procalcitonin (ng/mL) | 1.36 (0.18-9.97) | 0.76 |  |  |
| Ferritin ($\times 10^3$ng/mL) | 0.37 (0.10-1.47) | 0.16 | | |

|  |  |  |
| --- | --- | --- |
| D-dimer ($\mu$ g/mL) | 0.10 (0.96-1.03) | 0.82 |

Severity
|  |  |  |
| --- | --- | --- |
| 1 or 2 | (reference) | (reference ) |
| 3 | 1.52 (0.18-13.24) | 0.70 |
| 4 | 1.80 (0.14-23.37) | 0.65 |

We defined COVID-19 disease severity at administration as follows: severity level 1, hospitalized but not requiring supplemental oxygen; level 2, hospitalized and requiring supplemental oxygen ≤ 4 L per minute (L/min); severity level 3, hospitalized and requiring oxygen therapy ≥ 5 L/min or receiving nasal high-flow oxygen therapy, non-rebreather, or noninvasive mechanical ventilation; level 4, receiving invasive mechanical ventilation at administration.
BMI, body mass index; CI, Confidence interval; CRP, C-reactive protein; KL-6, Krebs von den Lungen-6; LDH, lactate dehydrogenase; OR, odds ratio.

### Factors contributing to improvement in respiratory status

In the univariate logistic regression analysis, factors contributing significantly to the improvement of respiratory status were BRT use, young age, absence of chronic heart disease, chronic kidney disease or hypertension, more than seven days from onset to drug administration, and use of any anti-viral drug (Table 3). However, in multivariate analysis, BRT use was not a contributing factor (OR = 1.75, 95 %CI 0.35 –8.67, P=0.50), while the use of the anti-viral drug was an independent contributing factor (OR =6.5, 95 %CI 1.13 – 37.56, P=0.04). Early administration of biological agents was the risk factor that reduced the likelihood of improving the respiratory status (OR = 0.82, 95 %CI 0.02 – 0.40, P=0.002).

**Table 3.**
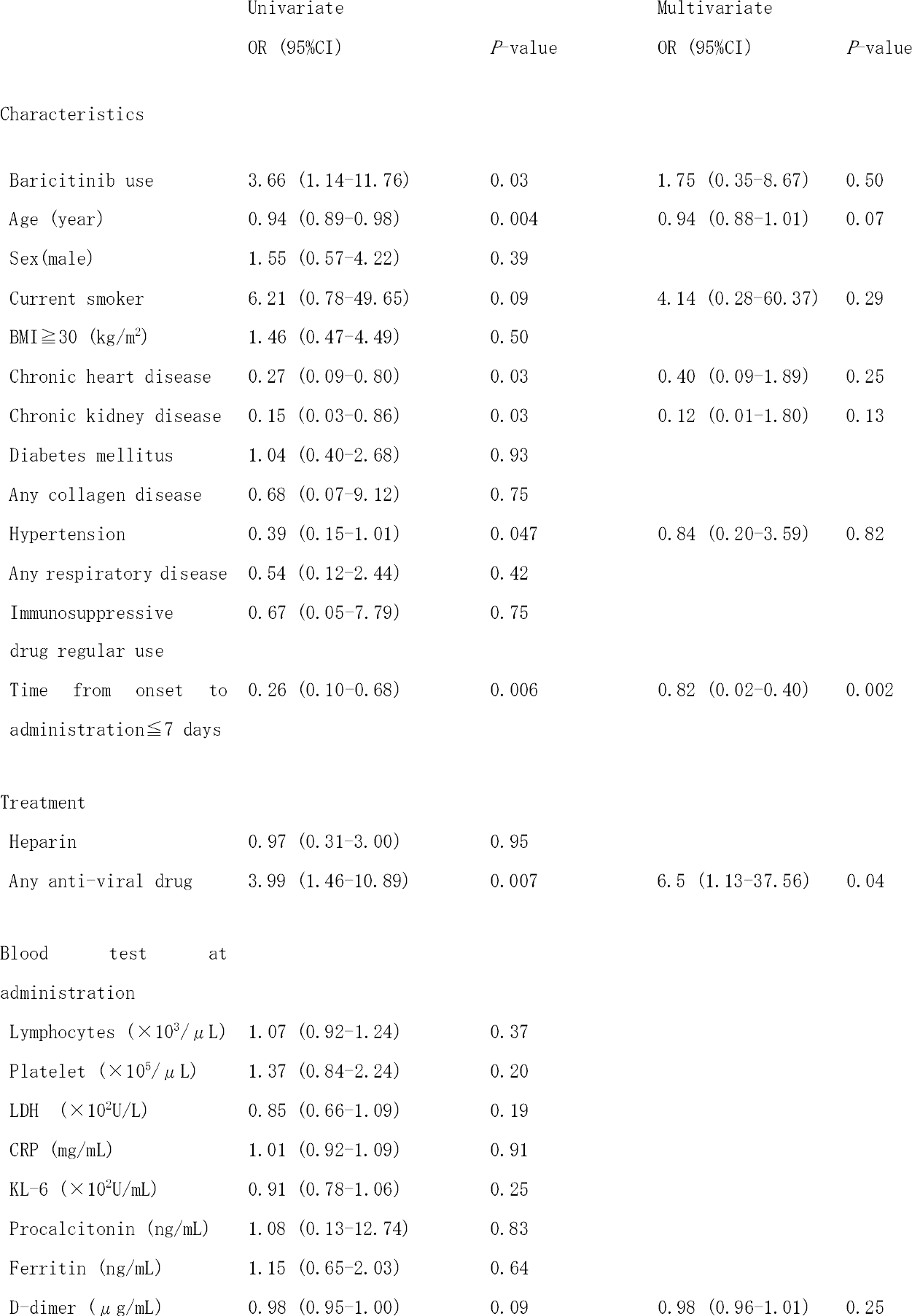
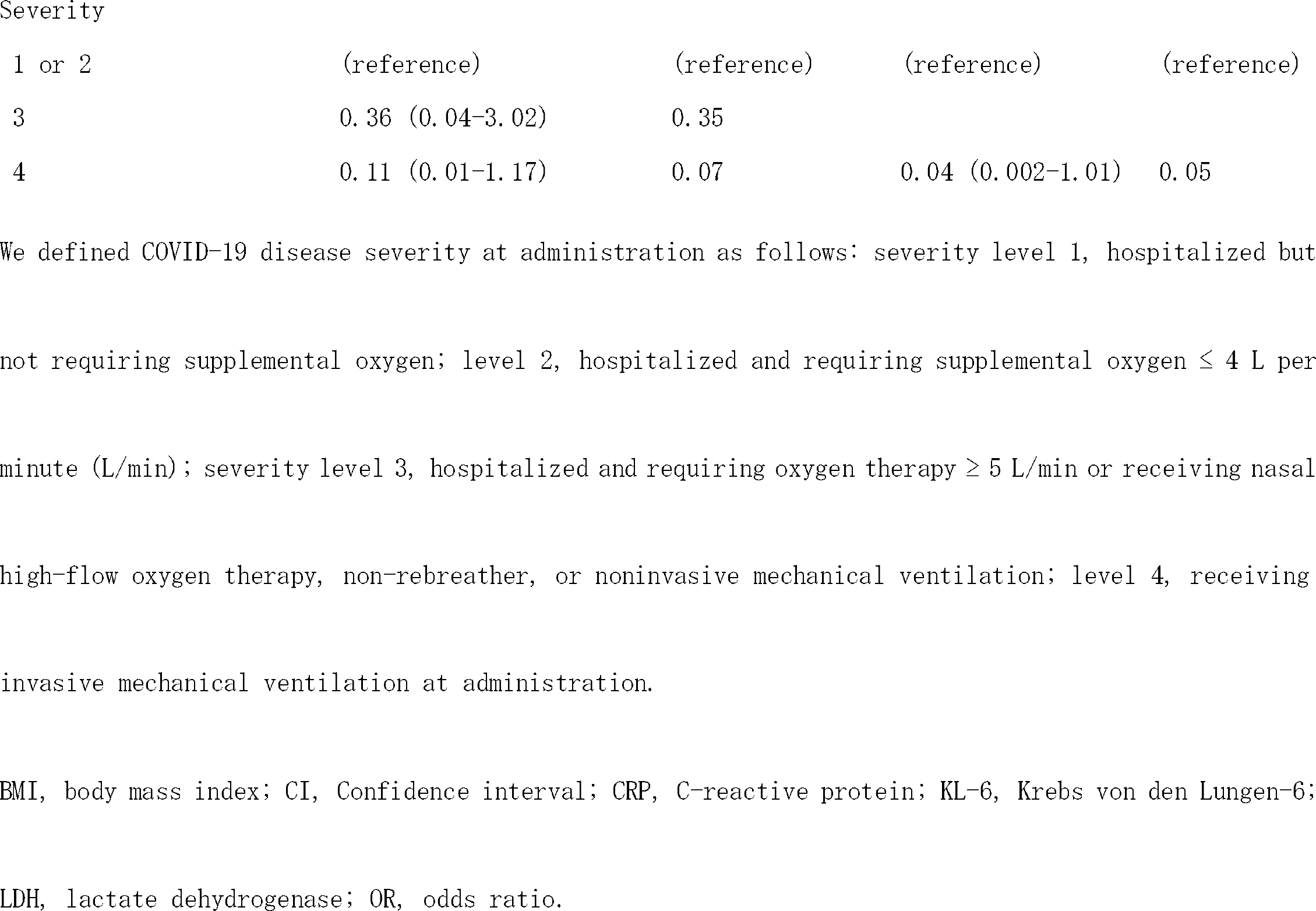
Predicting factors for improvement in respiratory status within 28 days of administration in patients treated with tocilizumab or baricitinib assessed using logistic regression analysis.

### Development of secondary infection

The rates of acquiring any secondary infection in patients within 28 days after initiation of treatment with TCZ and BRT were 15.6 and 14.7%, respectively. Univariate analysis did not identify any factors associated with the development of secondary infection after initiation of treatment with biological agents (Supplementary table). There was also no significant difference in infection-free survival (P=0.95) (Figure 2).

**Figure 2.**
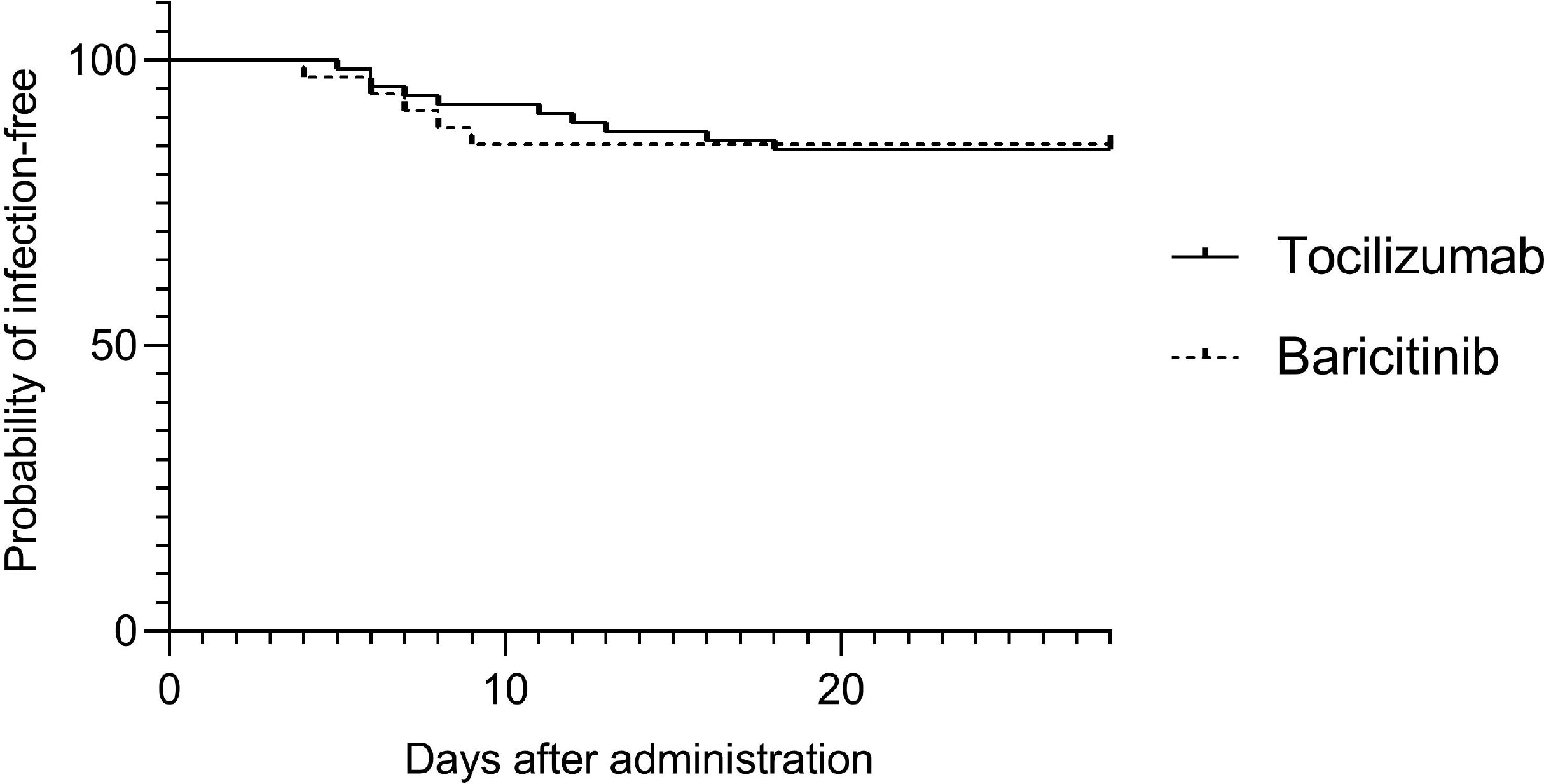
Kaplan–Meier estimates of infection-free survival between tocilizumab and baricitinib.

## Discussion

In this retrospective study, we compared the clinical characteristics between two groups of patients treated with TCZ and BRT for COVID-19. Multivariate analysis revealed that both the biological agents did not increase all-cause mortality within 28 days of treatment initiation. Age, underlying diseases, and early administration of biological agents were independent risk factors for all-cause mortality. None of the two biological agents significantly contributed to improving the respiratory status within 28 days. Use of anti-viral drugs and late administration of biological agents significantly contributed to improvement in respiratory status. No significant difference was observed in the development of secondary infection within 28 days after TCZ or BRT administration.

The results of this study showed that most of the patients who were treated with TCZ or BRT were also receiving steroid therapy. In addition, the severity of COVID-19 at the time of initiating treatment with biological agents was not significantly different between the two groups. In our hospital, patients with COVID-19 who required oxygen are usually treated with steroids, and biological agents are additionally administered to patients with increased oxygen demand, based on the guideline treatment plan (9, 10). Although the present study was retrospective, the strength of the study was that the baseline treatment and respiratory status of both groups are well matched.

According to the results of univariate analysis, TCZ seemed to increase the risk of 28-day mortality, while not improving the respiratory status. However, it was not identified as a significant risk factor in multivariate analysis. The reason for this could be due to the existence of multiple confounding factors for the use of TCZ. Comparison of clinical characteristics between the TCZ and BRT groups revealed an older median age and a higher proportion of patients with chronic diseases in the TCZ group, than in the BRT groups. Moreover, blood test results showed higher levels of LDH and D-dimer in the TCZ group than that in the BRT group. Advanced age and presence of underlying diseases are considered poor prognostic factors for COVID-19 (11, 12). Higher levels of LDH and D-dimer are associated with increased mortality in COVID-19 and are known to be predictors of severe disease (13). Although we did not find a significant difference in the severity of COVID-19 based on respiratory status, TCZ group might have potentially been at a higher risk for critical course. TCZ was shown to be effective relatively earlier than BRT in the COVID-19 pandemic and was used earlier in clinical practice. In contrast, BRT became widely used in Japan after approval for the treatment of COVID-19. In addition, backgrounds of the patients admitted to our hospital differed depending on the timing of spread of COVID-19. These circumstances might contribute to the bias in the clinical characteristics of the two groups.

Biological drugs are a risk factor for serious infection in rheumatoid arthritis (14). Although the COVID-19 trials showed no difference in the incidence of infections in either TCZ or BRT groups, compared to that in the placebo (4, 8), one retrospective study showed that concomitant use of TCZ and methylprednisolone is a risk factor for bacteremia (15). Besides, whether there is a difference in the risk of developing infections between TCZ and BRT has not been evaluated earlier. In our study, we found no difference in the incidence of secondary infection between TCZ group and BRT group. In addition, no risk factors were identified in the univariate analysis, which could be associated with the occurrence of secondary infection. Although we have not been able to verify whether the complications of infections had an impact on patient prognosis, neither biological drug seems to pose a significant risk of infection.

Given the fact that treatment with any of the two biological agents did not result in significantly different outcomes for patients, either TCZ or BRT can be selected in terms of efficacy and safety. BRT is an oral drug that can be administered even if the intravenous route is difficult to secure, and it is easy to discontinue. Tocilizumab is an intravenous or subcutaneous drug that can be used by patients who have difficulty with oral intake and in those with severe renal dysfunction. The choice should be based on the characteristics of each drug in each individual patient.

Our study confirmed that the improvement of patients’ respiratory status in COVID-19 was similar with both biological agents. In contrast, we found that the use of anti-viral drugs was significantly associated with improvement in respiratory status within 28 days.

According to the clinical trial, remdesivir has been shown to shorten the time to recovery in patients hospitalized with COVID-19 and with evidence of pneumonia (16). In contrast, several studies have failed to show clear efficacy (17-19); the efficacy of remdesivir as a single agent or adjunctive drug for standard care may be limited. Based on the results of our study, remdesivir may be an important drug that should essentially be administered to patients receiving biologic agents and steroids. This finding suggests the additive effect of remdesivir, which requires further investigation.

It is worth discussion that shorter time from the onset of illness to the administration of the biological drugs was an independent factor for poor prognosis. For both TCZ and BRT, earlier administration of drugs in the trials is associated with a greater reduction in the risk of death (5, 8). The reason of this apparently paradoxical observation was probably that the short time between the onset of symptoms and the administration of biological agents may reflect the rapid deterioration in respiratory status. In our hospital, biologic drugs are mostly administered to patients with increased oxygen demand and increased severity of illness. Worsening of respiratory status early in the course of the disease may be a prognostic factor that cancels out the benefit of early administration of biological drugs. The prognosis of patients who deteriorate rapidly after the onset of illness should be evaluated in future studies.

This study has several limitations. First, since the study was retrospective, prospective validation is needed to show the comparative efficacy of TCZ and BRT. Second, variant strains of SARS-CoV-2 and changes in healthcare availability that may affect patient outcomes were not validated in this study due to lack of data. Lastly, the efficacy of using the biological agents as a standalone treatment for COVID-19 was not verified in this study. However, the efficacy of TCZ and BRT has already been proven in previous studies. Our study was conducted to suggest a more favorable treatment based on these evidence-based practices of using TCZ and BRT.

In conclusion, the use of TCZ versus BRT had no different impact on all-cause mortality, improvement in respiratory status, and the development of secondary infection in patients diagnosed with COVID-19. In light of our findings, both biological agents are expected to be equally safe and clinically effective, although future prospective studies are needed.

## Data Availability

All data produced in the present study are available upon reasonable request to the authors

## Acknowledgements

Not applicable.

## Ethical approval

The research protocol was approved by the Ethics Committee of the Hokkaido University Hospital (Research No. 020-0107). The study was based on existing samples collected in the course of routine practice and no additional risks are posed to patients. Therefore, the individual’s informed consent was waived by the above ethics committee. Informed consent for study participation was officially announced on the website. All patient data were anonymized prior.

## Contributions

YK and SN contributed to the study concept and design and interpretation, statistical analysis, and drafting of the manuscript. KK, YY, NT, JN, MM, HH, KS and HS contributed to the acquisition and interpretation of data. NT contributed to statistical analysis. YK, SN, and MM contributed to data acquisition. MS and SK contributed to the study concept and design, acquisition and interpretation of data, and finalization of the manuscript. All authors read and approved the final manuscript.

## Abbreviations

BMI: body mass index
BRT: baricitinib
CI: Confidence interval
COVID-19: Coronavirus disease, 2019
CRP: C-reactive protein
IQR: interquartile rage
JAK: Janus kinase inhibitor
KL-6: Krebs von den Lungen-6
LDH: lactate dehydrogenase
OR: odds ratio
PCR: Polymerase chain reaction
SARS-CoV-2: severe acute respiratory syndrome coronavirus 2
TCZ: tocilizumab
WBC: White blood cell

