## Supplemental Table for "Comparative efficacy of tocilizumab and baricitinib in COVID-19 treatment: a retrospective cohort study"

**Supplementary table. Predicting risk factors for infection event within 28 days of administration in patients treated with tocilizumab or baricitinib assessed using logistic regression analysis.**

|  | univariate | |
| --- | --- | --- |
|  | OR (95%CI) | *P*-value |
| Characteristics |  |  |
| Baricitinib use | 0.93 (0.29-2.98) | 0.904 |
| Age (year) | 1.03 (0.98-1.08) | 0.208 |
| Sex(male) | 0.63 (0.19-2.08) | 0.453 |
| Current smoker | 0.75 (0.15-3.74) | 0.728 |
| BMI≧30 (kg/m^2^) | 0.69 (0.18-2.73) | 0.694 |
| Chronic heart disease | 2.96 (0.86-10.18) | 0.085 |
| Diabetes mellitus | 0.83 (0.26-2.68) | 0.767 |
| Hypertension | 2.61 (0.82-8.31) | 0.104 |
| Time from onset to administration≦7day | 2.43 (0.79-7.48) | 0.123 |
| Treatment |  |  |
| Heparin | 0.66 (0.18-2.33) | 0.657 |
| Any anti-viral drug | 0.55 (0.17-1.83) | 0.332 |
| Blood test at administration |  |  |
| Lymphocytes (×10^3^/μL) | 0.97 (0.82-1.15) | 0.747 |
| Platelet (×10^5^/μL) | 0.88 (0.50-1.56) | 0.672 |
| LDH (×10^2^U/L) | 1.10(0.82-1.49) | 0.498 |
| CRP (mg/mL) | 1.00 (0.90-1.10) | 0.897 |
| KL-6 (×10^2^U/mL) | 1.08 (0.91-1.29) | 0.373 |
| Procalcitonin (ng/mL) | 1.22 (0.17-8.46) | 0.844 |
| Ferritin (ng/mL) | 1.11 (0.65-1.88) | 0.706 |
| D-dimer (μg/mL) | 1.02 (1.00-1.04) | 0.060 |

BMI, body mass index; CI, Confidence interval; CRP, C-reactive protein; KL-6, Krebs von den Lungen-6; LDH, lactate dehydrogenase; OR, odds ratio.
